# Population-based sero-epidemiological estimates of real-world vaccine effectiveness against Omicron infection in an infection-naive population, Hong Kong, January to July 2022

**DOI:** 10.1101/2022.11.01.22281746

**Authors:** Jonathan J Lau, Samuel MS Cheng, Kathy Leung, Cheuk Kwong Lee, Asmaa Hachim, Leo CH Tsang, Kenny WH Yam, Sara Chaothai, Kelvin KH Kwan, Zacary YH Chai, Tiffany HK Lo, Masashi Mori, Chao Wu, Sophie Valkenburg, Gaya K Amarasinghe, Eric HY Lau, David S Hui, Gabriel M Leung, Malik Peiris, Joseph T Wu

## Abstract

The SARS-CoV-2 Omicron variant has demonstrated enhanced transmissibility and escape of vaccine-derived immunity. While current vaccines remain effective against severe disease and death, robust evidence on vaccine effectiveness (VE) against all Omicron infections (i.e. irrespective of symptoms) remains sparse. We addressed this knowledge-gap using a community-wide serosurvey with 5,310 subjects by estimating how vaccination histories modulated risk of infection in Hong Kong (which was largely infection naïve) during a large wave of Omicron epidemic during January-July 2022. We estimated that Omicron infected 45% (41-48%) of the Hong Kong population. Three and four doses of BNT162b2 or CoronaVac were effective against Omicron infection (VE of 47% (95% credible interval 34-68%) and 70% (43-99%) for three and four doses of BNT162b2 respectively; VE of 31% (1-73%) and 59% (10-99%) for three and four doses of CoronaVac respectively) seven days after vaccination, but protection waned with half-lives of 15 (3-47) weeks for BNT162b2 and 5 (1-37) weeks for CoronaVac. Our findings suggest that booster vaccination can temporarily enhance population immunity ahead of anticipated waves of infections.

## Main text

During 1 January - 31 July 2022, Hong Kong experienced an unprecedented fifth wave of COVID-19 infections driven predominantly by the Omicron BA.2 variant (B.1.1.529.2) with 1,341,363 reported cases (18.4% of the population) and 9,290 deaths (0.7%) [1]. The fifth wave dwarfed the previous four waves in terms of cumulative infection attack rate (IAR), which was nearly zero before 2022 given Hong Kong’s then successful “dynamic Zero-Covid” strategy. Thus, population immunity to SARS-CoV-2 was almost entirely vaccine-derived when the fifth wave began. The mRNA vaccine Comirnaty (BNT162b2 mRNA, BioNTech/Fosun-Pharma, Mainz, Germany/Shanghai, China) and the inactivated CoronaVac vaccine (Sinovac Life Sciences, Beijing, China) have been available free of charge to Hong Kong residents since 26 February 2021. Population uptake of at least two doses of either vaccine increased from 4.7 million (66%) on 1 January to 6.5 million (93%) on 31 July [1].

We conducted a serial cross-sectional serosurvey to estimate: (i) age-specific IAR in the fifth wave; and (ii) vaccine effectiveness (VE) against SARS-CoV-2 infection conferred by two, three and four homologous doses of BNT162b2 or CoronaVac. Specifically, for each subject in our serosurvey, we estimated the probability of being infected by SARS-CoV-2 before study recruitment, given age, vaccination record and seropositivity of the serum sample (see Methods). We assumed that: (i) daily age-specific force of infection was proportional to daily viral load from city-wide wastewater surveillance (see **Figure 1**) which has been shown to be a robust (normalized) proxy for disease prevalence over time [2-4]; and (ii) one-dose vaccination provided no protection against infection and each successive homologous dose conferred greater VE which decayed exponentially over time at the same rate between doses of the same vaccine [5].

**Figure 1:**
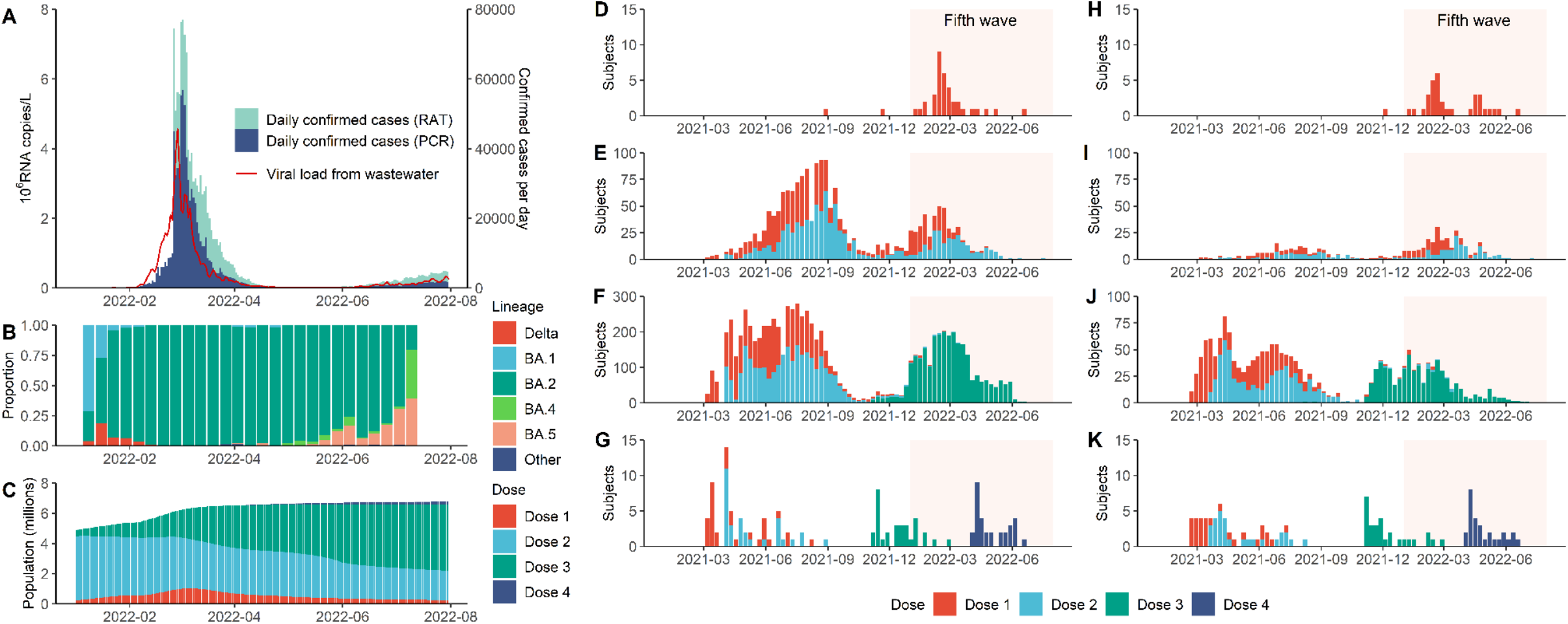
Daily COVID-19 confirmed cases, SARS-CoV-2 wastewater viral load, weekly proportion of SARS-CoV-2 lineages, population vaccination coverage and study subject vaccination history. **A**. Daily confirmed COVID-19 cases in Hong Kong stratified by cases confirmed by RT-PCR (PCR) or rapid antigen testing (RAT), superimposed against two-day running geometric mean viral load per capita (in millions of copies of SARS-Cov-2 RNA/L) detected in city-wide wastewater surveillance. **B**. Weekly proportion of SARS-CoV-2 lineages detected in Hong Kong via genome sequencing as uploaded to GISAID by 14 September 2022. **C**. Total population with one to four doses of BNT162b2 and/or CoronaVac in Hong Kong. **D.-G**. Vaccination history for study participants with one (**D**), two (**E**), three (**F)** and four (**G**) homologous doses of BNT162b2 at the time of sample collection. Shaded area corresponds to the duration of the fifth wave from 1 January to 31 July 2022. **H.-K**. same as **D-G** for CoronaVac.

Our serosurvey subjects included: (i) 5,173 healthy adult blood donors recruited from the Hong Kong Red Cross Blood Transfusion Service between 28 April and 30 July 2022; and (ii) 137 children aged 18 months to 11 years randomly recruited from the community to participate in an independent polio sero-epidemiology study. Vaccination histories were available for 5,242 subjects (see **Figure 1** and **Extended Data Table 1**) from the Hong Kong Department of Health (98%) or self-report (2%). At the time of sample collection, 1,237 blood donors (24%) and 31 children subjects (23%) self-reported a previous infection.

We developed two in-house ELISA assays detecting IgG antibodies to the C-terminal domain of the nucleocapsid (N) protein (N-CTD) and the Open Reading Frame 8 protein (ORF8) of SARS-CoV-2, respectively, modifying and validating previously reported methods[6, 7]. We estimated that our N-CTD assay was more than 95% sensitive and 96% specific in detecting recent Omicron infection among unvaccinated individuals and homologous BNT162b2 vaccinees. Because the inactivated whole-virus vaccine CoronaVac elicits antibody to the N protein, the ORF8 assay was optimized specifically for discriminating between infection and vaccine-derived antibody in CoronaVac vaccinees. We estimated that our ORF-8 assay was 81% sensitive and 93% specific in detecting recent Omicron infection among homologous CoronaVac vaccinees. See **Extended Data Figure 2** for a mapping of vaccination cohort by assay. See Methods and **Extended Data Figures 3 and 4** for further details on assay workflow, performance and output. To our knowledge, our ORF8 assay is the first serological test that could effectively detect recent and discriminate SARS-CoV-2 infection from vaccination among CoronaVac vaccinees.

**Figure 2:**
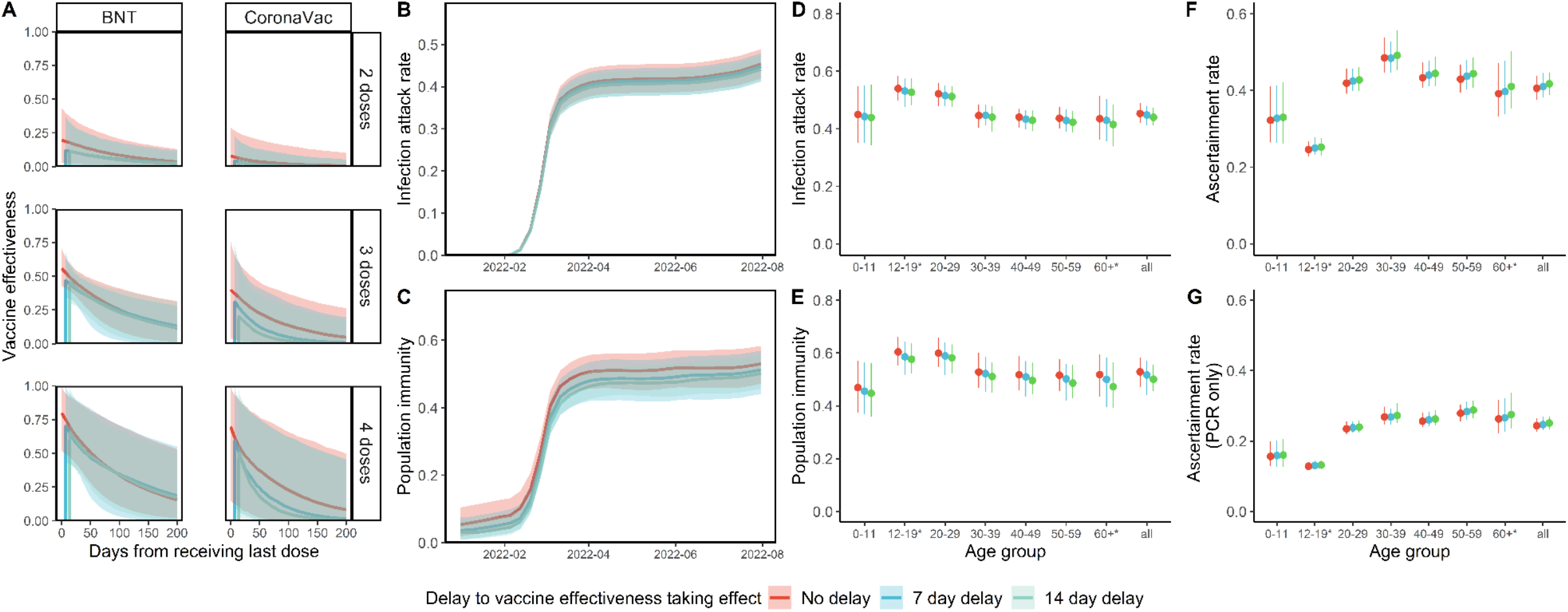
Estimated vaccine effectiveness (VE), infection attack rate (IAR), population immunity and ascertainment ratio. (**A**.) VE at zero to 200 days from receipt of last dose. VE is presented separately over time for two, three or four homologous doses of BNT162b2 (BNT) or CoronaVac. (**B**.) IAR over time, (**C**.) population immunity over time from infection and vaccination, (**D**.) cumulative IAR by 31 July 2022, (**E**.) cumulative population immunity by 31 July 2022 and (**F.-G**.) ascertainment rate based on all cumulative confirmed cases (**F**.) or cumulative RT-PCR confirmed cases only (**G**.). All analyses were stratified by assumptions on delay to VE taking effect and (**D.-G**. only) age group. (**A.-C**. only) The lines indicate posterior medians and shaded bars indicate 95% credible intervals based on the fitted model. (**D.-G**. only) The dots indicate posterior medians and the lines indicate 95% credible intervals based on the fitted model. * Cumulative IAR, population immunity and ascertainment rate estimates among those aged 12-19 or aged 60 or above were less accurate as no subjects were between 12 and 17 years old, and few were above 65 years old.

Assuming VE took full effect seven days after vaccination, we estimated (i) VE for the 2^nd^, 3^rd^ and 4^th^ doses of BNT162b2 were 12% (95% credible interval: 1-37%), 47% (34-68%) and 70% (43-99%) seven days following immunization, respectively, waning with half-life of 104 (20-328) days (i.e. 3^rd^- and 4^th^-dose VE declined to 25% (2-37%) and 35% (3-71%) 100 days after immunization); and (ii) VE for the 2^nd^, 3^rd^ or 4^th^ dose of the CoronaVac vaccine were 4% (0-23%), 31% (1-73%) and 59% (10-99%) seven days following immunization, respectively, waning with half-life of 37 (10-258) days (i.e. 3^rd^- and 4^th^-dose VE declined to 5% (0-31%) and 10% (0-62%) 100 days after immunization). Sensitivity analyses assuming VE takes effect instantaneously after vaccination or after a fourteen-day delay yielded similar results (**Figure 2**).

Using our VE estimates and (anonymized) vaccination records for every individual in the population provided by the Hong Kong government, we estimated that SARS-CoV-2 (predominantly Omicron BA.2) infected 45% (41-48%) of the population between 1 January and 31 July, 2022. Adolescents and young adults had slightly higher IARs than overall. Population immunity (conferred by both infection and vaccination) reached 52% (44-57%) by 31 July 2022. Assuming VE takes effect instantaneously or after a fourteen-day delay yielded similar estimates. Overall ascertainment ratio was 25% (23-27%) from reverse transcription polymerase chain reaction (RT-PCR) testing alone, increasing to 41% (38-45%) if augmented with rapid antigen testing (RAT) (**Figure 2**). Our Omicron IAR estimates were lower than those reported in South Africa (58% in urban areas)[8], Denmark (66%)[9] and Navarre, Spain (59%)[10], likely reflecting the effectiveness of extensive public health and social measures (PHSMs) imposed in Hong Kong during the fifth wave, such as an universal mask mandate with high community compliance, closure of all bars and limits on opening hours and new ventilation requirements in all restaurants.

Estimating VE against Omicron infection has been challenging in populations that have experienced widespread infection by older variants, due to difficulties in disentangling the protective effect of vaccine-derived immunity from that of prior infection-derived immunity and hybrid immunity. While the test-negative design has been increasingly used to estimate VE against Covid-19, the robustness of the resulting estimates are typically conditional on symptoms and susceptible to confounding and selection bias (e.g. due to differential healthcare seeking behaviour) [11]. Furthermore, most VE estimates hitherto have estimated protection against symptomatic disease, hospitalization or death but not against all infections including asymptomatic infections. Our estimates of VE against Omicron infection are robust against the abovementioned limitations because Hong Kong had negligible infection-derived immunity against any SARS-CoV-2 prior to January 2022 and the infection history of our subjects were individually inferred based on their serological measurements (irrespective of history of symptoms, case confirmation or contact) [12].

Our estimates provide evidence of the short-term effectiveness against Omicron infection of a third or fourth dose of either the mRNA or inactivated vaccine. Slightly higher initial BNT162b2 VE followed by rapid waning has been reported for symptomatic Omicron BA.2 infection. For example, Chemaitelly et al. reported effectiveness against symptomatic infection after the second dose was 51.7% in the first three months and waned to ≤10% thereafter, increasing to 43.7% after a booster dose before waning again at similar rate[13]. Meanwhile, Gazit et al. reported a fourth dose of BNT162b2 was 65.1% more effective by the third week against RT-PCR confirmed Omicron infection relative to a third dose, declining to 22.0% by end of ten weeks[14], although lower effectiveness was reported in separate Israeli studies [15, 16]. In contrast, there is very limited data on CoronaVac VE against Omicron infection [17]. Our study provides the first estimate of real-world VE and waning against Omicron infection conferred by three or four doses of CoronaVac. A recent telephone survey in Hong Kong reported three doses of COVID-19 vaccination with either BNT162b2 or CoronaVac provided 52% protection against test-positivity by PCR or rapid antigen testing (RAT) but was unable to account for time since vaccination or for asymptomatic infection [18]. Two South American studies reported 38.2% and 39.8% VE from two doses of CoronaVac against symptomatic Omicron infection in children aged 3-5 and 6-11 years respectively[17, 19].

We previously reported markedly reduced serum neutralizing antibody titers against BA.2 amongst individuals recently vaccinated with three doses of CoronaVac as compared to the wildtype virus, with antibody titres below the predicted protective threshold [20]. Thus, our estimate of VE against BA.2 infection elicited by three doses of CoronaVac appears greater than would be expected from neutralizing antibody titres. Indeed, a recent VE study of CoronaVac vaccine using a test-negative design during this same BA.2 epidemic in Hong Kong also observed significant protection from severe disease and death [21]. It is possible that neutralizing antibody titres under-estimate protection conferred by whole virus inactivated vaccines such as CoronaVac, which present multiple viral proteins to the host immune system that may protect via multiple pathways other than neutralizing antibodies, such as T-cell immunity and antibody dependent cytotoxicity [6].

Our study has several important limitations. First, we assumed that the effect of vaccination history on contact patterns and mobility (i.e. exposure to the virus), a potential confounder of VE estimates, was negligible. Second, we had no serum samples from individuals aged 12-17 and only few samples from individuals aged >65 years. As such, our IAR estimates for these age groups were less robust compared to that for other age groups. Third, our analysis was primarily based on seroprevalence amongst blood donors who might not be representative of the general population in terms of their infection history. Nonetheless, we previously observed similar influenza seroprevalence between blood donors, hospital outpatients and other community subjects during the 2009 influenza pandemic [22]. Given the high transmissibility of Omicron, we posit that seroprevalence may also be similar between blood donors and the general population. Fourth, the small number of CoronaVac vaccinees in our serosurvey together with the short duration of the fifth wave led to substantial uncertainty in our CoronaVac VE estimates. Fifth, we were unable to estimate VE conferred by heterologous vaccinations (CoronaVac with BNT boosters or vice versa) due to the very small number of individuals with such vaccination history (heterologous boosters were not available in Hong Kong until late 2021). When estimating IAR, we assumed that VE for each dose in heterogenous vaccinees equals that of the corresponding dose in homologous vaccinees. Sixth, as our positive controls comprise only of confirmed or self-reported infections, the corresponding seropositivity threshold may not be sufficiently sensitive to detect individuals with asymptomatic or very mild infections, thereby underestimating IAR.

In conclusion, our results indicate the short-term effectiveness of booster vaccination using either the mRNA or inactivated vaccine in preventing SARS-CoV-2 Omicron BA.2 infection. As such, surge booster campaigns could be strategically used to rapidly boost population immunity prior to upcoming waves of infections. For example, a preparatory surge booster campaign in mainland China may temporarily mitigate the anticipated wave of infections resultant from exiting its dynamic Zero-Covid strategy. The comparatively lower infection attack rate in Hong Kong also highlights the effect of supplementing vaccination campaigns with continued PHSMs in disease transmission. Nonetheless, in light of antigenic imprinting, rapid viral evolution and debut of new or updated COVID-19 vaccines, more studies quantifying the protective effect of repeated booster vaccination are necessary for policymakers to develop effective booster vaccination strategies.

## Data Availability

The anonymized vaccination record data were compiled by the Office of the Government Chief Information Officer (OGCIO) and the Department of Health, The Government of Hong Kong Special Administrative Region (HKSAR). Age data for confirmed cases were compiled by the Centre for Health Protection. Data on viral load from sewage surveillance were compiled by the Environmental Protection Department, The Government of HKSAR. The aforementioned data could not be shared due to confidentiality undertakings to the above-named agencies. Interested parties could contact these agencies for access to these data.
Anonymized serology output data are available from the authors upon request. Outputs of our analysis and other source data are accessible at https://github.com/jonathanjlau- hku/hkserosurvey2022.

## Online Methods

### Data sources

#### Serosurveys conducted by the study team

As part of a community-based COVID-19 sero-epidemiological study, we recruited healthy blood donors by convenience sampling at the five largest blood donation centres (Mongkok, Causeway, Kwun Tong, Tsuen Wan and Shatin) of the Hong Kong Red Cross Blood Transfusion Service (HKRCBTS) from 28 April 2022 to 30 July 2022. We also tested serum samples from participants of an independent polio sero-epidemiology study targeting children aged 18 months to 10 years from 7 May to 5 August 2022. Blood donors were matched by the HKRCBTS and the Hong Kong Department of Health with official vaccination records via unique Blood Transfusion Service donor identification numbers. The records were then anonymised and provided to the study team. Both blood donors and children participants were asked to self-report their vaccination and COVID-19 infection history. In cases where official vaccination records were unavailable (i.e. those vaccinated outside Hong Kong), we relied on the donors’ self-reported vaccination history if provided. All children participants self-reported their vaccination and infection history.

Written informed consent was obtained from all participants. Parental consent was obtained for all participants aged < 18. Ethical approval for this study was obtained from the Institutional Review Board of the Hospital Authority Hong Kong West Cluster / University of Hong Kong (IRB No. UW 20-132).

#### Vaccination records, confirmed cases and sewage surveillance data provided by the Hong Kong government

Official vaccination records in Hong Kong are maintained by the Hong Kong Department of Health (DH) [23]. Anonymous data on every vaccination up to July 31, 2022, including the date of each dose, type of vaccine (BNT162b2 or CoronaVac) used and vaccinee year-of-birth, were compiled by DH and provided to us by the Hong Kong Office of the Government Chief Information Officer (OGCIO). Age data on all confirmed SARS-CoV-2 cases were provided by the Centre for Health Protection. Daily per-capita two-day running geometric mean SARS-CoV-2 viral load data (in copies of SARS-CoV-2 RNA/L) obtained from city-wide COVID-19 wastewater surveillance up to July 31, 2022 were provided by the Hong Kong Environmental Protection Department. 2022 projected mid-year population in each age cohort was obtained from the Hong Kong Census and Statistics Department [24].

### Laboratory methods

We developed two in-house ELISA assays which detected IgG antibodies to the C-terminal domain of the nucleocapsid (N) protein (N-CTD) and the Open Reading Frame 8 protein (ORF8)[25] of SARS-CoV-2, respectively, modifying the methodology reported in Mok et al.[6] and Hachim et al[7]. The ORF8 assay was developed specifically for detecting past Omicron BA.2 infections in CoronaVac vaccinees because most of them were N-CTD-seropositive due to the immune response that CoronaVac elicits against the N protein. The ELISA assays as previously described[6, 7] were optimised and validated. In brief, 96-well ELISA plates (Nunc MaxiSorp, Thermo Fisher Scientific) were coated overnight with 40 ng/well purified recombinant N-CTD protein in PBS buffer for N-CTD protein ELISA, 30 ng/well purified recombinant ORF8 protein in PBS buffer for ORF8 ELISA. The plates were then blocked by 100 μl of Chonblock blocking buffer (Chondrex Inc, Redmond, US) per well, and were incubated at room temperature for 2 h. Each serum sample was tested at a dilution of 1:100 in Chonblock blocking buffer in duplicate. The serum dilutions were added and were incubated for 2 h at 37 °C. After extensive washing with PBS containing 0.2% Tween 20, horseradish peroxidase (HRP)-conjugated goat anti-human IgG (1:5,000, GE Healthcare) was added and incubated for 1 h at 37 °C. The ELISA plates were then washed again with PBS containing 0.2% Tween 20. Subsequently, 100 μL of HRP substrate (Ncm TMB One; New Cell and Molecular Biotech Co. Ltd, Suzhou, China) was added into each well. After 15 min incubation, the reaction was stopped by adding 50 μL of 2 M H2SO4 solution and analyzed on a microplate reader at 450 nm wavelength. Positive and negative controls were included in each run.

This resulted in cut-offs of 0.2583 and 0.33 optical density (OD) for N-CTD and ORF8 respectively. The assays and cut-offs were validated against pre-pandemic blood donor samples, blood samples from homologous BNT162b2 or CoronaVac vaccinated individuals collected during periods of minimal community transmission in 2020 and 2021, blood samples collected from RT-PCR confirmed SARS-CoV-2 convalescent individuals in 2020 and 2021, and samples from blood donors in the present study with self-reported infection history. See **Extended Data Figure 3** for details on control groups and assay performance (sensitivity, specificity and Receiver Operating Curves). We set the ELISA cut-offs to approximately maximize the sum of sensitivity and specificity, which were in turn estimated via bootstrapping 2,000 samples using the pROC R package[26], with specificity set not lower than 90%.

We did not account for waning of N-CTD and ORF8 antibody response. Nonetheless, we previously reported that N and ORF8 specific antibody responses were well maintained for at least 100 days post-infection[7]. This is on par with the time elapsed between infection (e.g. the the fifth wave peaked in early-March 2022) and the time of sample collection (between late April and July 2022) for our serosurvey subjects.

### Statistical methods

#### Statistical inference of VE

Let *VE*_*ν,j*_(*u*) be the VE of vaccine type *ν* (*B* for BNT162b2 and *C* for CoronaVac) against infection *u* days after the *j*th dose in a homologous series has taken effect. For each vaccine type *ν*, we assumed: (i) the first dose provided no protection against infection, i.e. *VE*_*ν*,1_ (*u*) = 0[27]; (ii) *VE*_*ν,j*_(0) depended on the number but not the time of previous doses; (iii) VE waned exponentially at rate *λ*_*ν*_ after each dose [5, 28, 29], i.e. *VE*_*ν,j*_(*u*) = *VE*_*ν,j*_(0) · exp(−*λ*_*ν*_*u*); and *VE*_*ν,j*_(0) increased with each successive dose in a homologous series, i.e. *VE*_*ν,j*+1_(0) > *VE*_*ν,j*_(0) [15, 30-32]). We also assumed that the initial VE of 2-dose BNT162b2 was not inferior to that of 2-dose CoronaVac, i.e. *VE*_*B*,2_(0) ≥ *VE*_*C*,2_(0) (the latter was not statistically identifiable otherwise).

Let time 0 be 1 January 2021. We assumed that the force of infection (FOI) at time *t* was proportional to the viral load per capita from city-wide sewage. Specifically, given an individual aged *a* with vaccination history H who remained uninfected at time *t*, her FOI at that time was

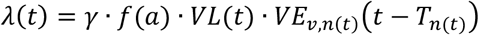

where:

1. *f*(*a*) was the effect of age on FOI with *f*(35) = 1 (i.e. those aged 35 years was the reference group). We assumed that: (i) *f*(*a*) was a piecewise cubic Hermite interpolating polynomial function for 10 ≤ *a* ≤ 65 with knots at 10, 18, 35, 50 and 65 years; and (ii) *f*(*a*) = *f*(10) for *a* < 10 and *f*(*a*) = *f*(65) for *a* > 65.
2. *VL*(*t*) was the two-day running geometric mean viral load per capita from city-wide sewage.
3. *n*(*t*) was the total number of doses of vaccine type *v* that the individual had received up to time *t* and *T*_n(*t*)_ was the time at which the most recent dose took effect.
4. *γ* was a scaling factor (subject to statistical inference; see below).

The probability that this individual was infected between time 0 and *t* was 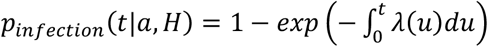. If tested at the time *t*, this individual would be seropositive with probability *P*_*seropositive*_(*t*|*a,H*) = *q*_*sens, ν*_ · *P*_*infection*_(*t*|*a,H*) + (1 − *q*_*spec,ν*_)· (1 − *P*_*infection*_ (*t*|*a,H*)) where *q*_*sens, ν*_ and *q*_*spec,ν*_ were the sensitivity and specificity of the serological assay that we used to infer previous Omicron infections for individuals vaccinated with vaccine type *ν*.

Let ***θ*** denote the set of model parameters subject to statistical inference (Table 1). Let **D** denote the data available for inferring ***θ*** which comprised:

1. The age, vaccination history and time of serum collection of each subject *i* in the serosurvey. These data were used to calculate the probability of seropositivity of the serum sample collected from subject *i* (*P*_*seropositive*.*i*_) via the abovementioned model.
2. The observed seropositivity of the serum sample for each subject *i* in the serosurvey (*τ* _*i*_ = 1 if seropositive and *τ* _*i*_ = 0 otherwise);
3. The number of positive and negative controls for estimating the sensitivity and specificity of our in-house ELISA assays among individuals with different vaccination history (*n*_*sens, ν*_ and *n*_*spec, ν*_) and the respective number of seropositive samples (*y*_*sens, ν*_ and *y*_*spec, ν*_). See **Extended Data Figure 3** for details.

**Table 1.** Parameters subject to statistical inference.

| Parameters | Description |
| --- | --- |
| $VE_{v,j}(0)$ for $v \in \{B, C\}, j \in \{2,3,4\}$ | Initial vaccine effectiveness conferred by each successive dose of vaccine |
| $\lambda_v$ for $v = \{B, C\}$ | Waning rate of VE |
| $\gamma$ | The scaling factor that related FOI to sewage viral load |
| $f(10), f(18), f(50)$ and $f(65)$ | The effect of age on FOI for age 10, 18, 50 and 65 years |
| $q_{sens,v}$ and $q_{spec,v}$ for $v \in \{U, B, C\}$ | Sensitivity and specificity of serological assays for detecting recent Omicron infection among unvaccinated individuals ($U$) and those vaccinated with BNT162b2 ($B$) or Coronavac ($C$) |

We used the following likelihood function to infer ***θ*** from **D**:

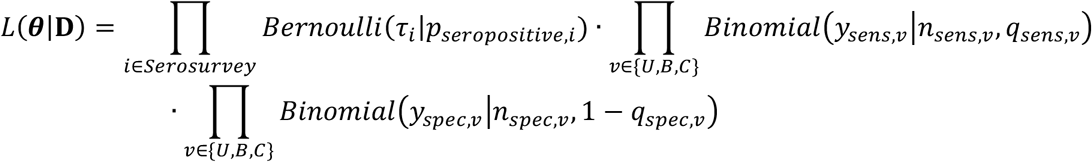

where (*Bernoulli* (·|*p*) was the Bernoulli pdf with parameter *p, Binomial* (· |*n,q*) was the Binomial pdf with *n* trials and success probability *q*. The statistical inference was performed in a Bayesian framework with noninformative (flat) priors using Markov Chain Monte Carlo with Gibbs sampling. We used *p*(***θ***) to denote the posterior distribution of ***θ*** obtained from fitting the model to the data **D**.

#### Estimating infection attack rate and population immunity

We randomly drew 300 samples of **θ** from *p*(***θ***). For each sample of ***θ*** drawn, we calculated *P*_*infection*.*i*_ (*t*) (probability of infection) and *P*_*infection,i*_(*t*) + (1 − *P*_*infection,i*_ (*t*)) · *VE*_*ν,j*_(*t*) (expected immunity) for each individual in the general population given her vaccination record (as done for our serosurvey subjects) at days = on weekly-intervals between 1 January and 31 July, 2022. Posterior medians and 95% credible intervals of age-specific IARs and population immunity were compiled accordingly.

For individuals with heterologous CoronaVac and BNT162b2 vaccinations, we assumed VE for each dose was the same as that of the corresponding type and dose in a homologous series. We substituted missing records for intervening or preceding doses with the vaccine type of the next recorded dose, with a 90-day gap between the third and fourth doses, 180-day gap between the second and third doses or a 14-day gap between the first and second doses as per Hong Kong government recommendations before 31 May 2022. We derived the number of unvaccinated individuals in each age cohort based on the 2022 predicted mid-year population per the Census and Statistics Department [24].

Lastly, we calculated the median and 95% confidence intervals of IARs, population immunity and ascertainment ratios by age-group (**Figure 2** and **Extended Data Figures 5 and 6)**. We further performed sensitivity analyses incorporating the posterior distribution corresponding to no (0-days) or a two-week (14-days) delay for VE to take effect after each dose (**Figure 2**).

All analyses were performed using MATLAB 2022a with the Parallel Computing and Econometrics toolboxes and R 4.2.1, with the tidyverse, pROC and cowplot packages.

## Data availability

The anonymized vaccination record data were compiled by the Office of the Government Chief Information Officer (OGCIO) and the Department of Health, The Government of Hong Kong Special Administrative Region (HKSAR). Age data for confirmed cases were compiled by the Centre for Health Protection. Data on viral load from sewage surveillance were compiled by the Environmental Protection Department, The Government of HKSAR. The aforementioned data could not be shared due to confidentiality undertakings to the above-named agencies. Interested parties could contact these agencies for access to these data.

Anonymized serology output data are available from the authors upon request. Outputs of our analysis and other source data are accessible at https://github.com/jonathanjlau-hku/hkserosurvey2022.

## Code availability

All code files are accessible at https://github.com/jonathanjlau-hku/hkserosurvey2022.

## Acknowledgements

We thank the following agencies from The Government of Hong Kong Special Administrative Region for compiling the data used in this research: Office of the Government Chief Information Officer (OGCIO), Department of Health, Centre for Health Protection and the Environmental Protection Department (see Data Availability for details). The study was supported by the Health and Medical Research Fund - Commissioned Research on the Novel Coronavirus Disease (COVID-19) (reference nos. COVID190126) from the Health Bureau, by AIR*a*InnoHK administered by Innovation and Technology Commission, both of the Government of the Hong Kong Special Administrative Region, and the Theme based Research Grants Scheme (T11-712/19-N, SAV) of the Hong Kong Special Administrative Region. JJ Lau is supported by the University of Hong Kong Presidential PhD Scholarship. The authors thank Dr. Jennifer Leung, Sr. Chelly Chu and Sr. Cathy Chan from the Hong Kong Red Cross Blood Transfusion Service, and Dr. Tommy Lam, Dominic Chan, Hermione Kock, Stephy Lam, Melanie Ngai, Zoe Song, Miky Wong and Nicole Wong from the School of Public Health, The University of Hong Kong, for research support.

