## Supplemental files for "Population-based sero-epidemiological estimates of real-world vaccine effectiveness against Omicron infection in an infection-naive population, Hong Kong, January to July 2022"

**Extended data: Figures**

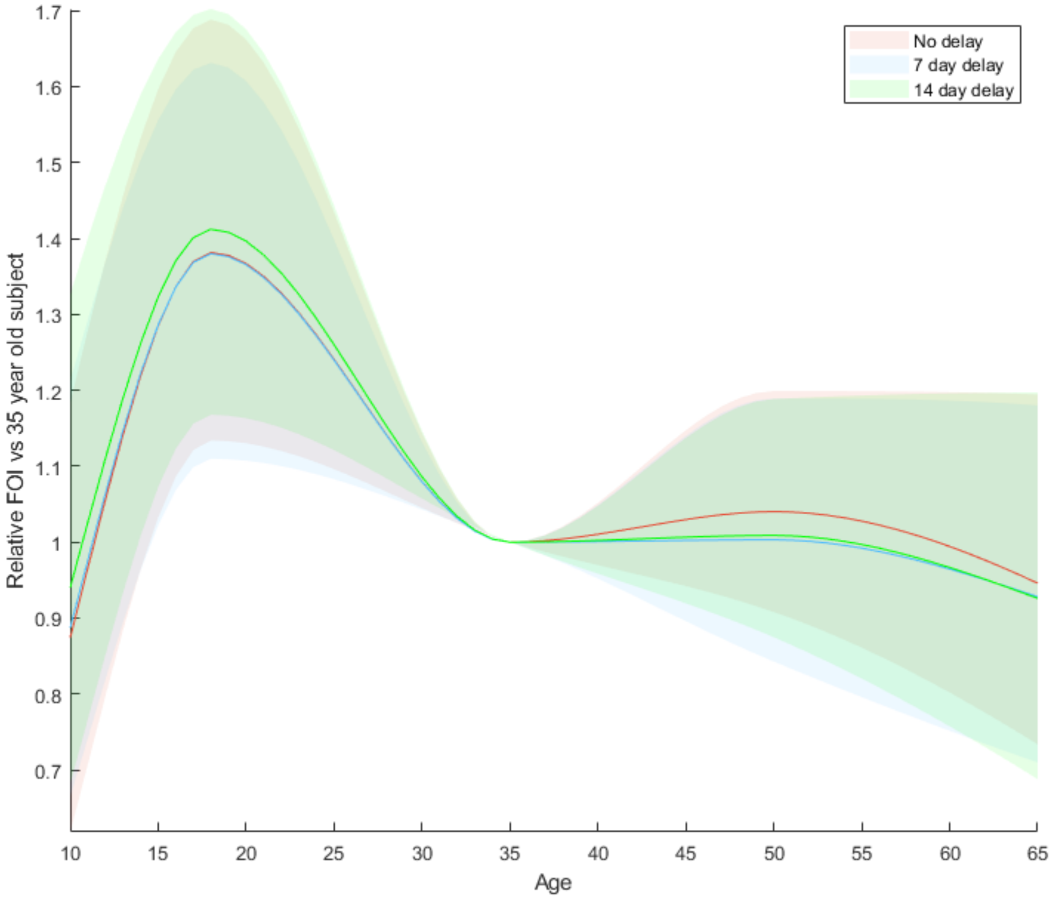


**Extended Data Figure 1: The effect of age on force of infection (FOI) (i.e. $f(a)$ in the model).** Individuals aged 35 years served as the reference group. The solid line indicates the posterior median, and the shaded regions indicate the 95% credible interval based on the fitted model, differentiated by assumption on delay to vaccine effectiveness taking effect.

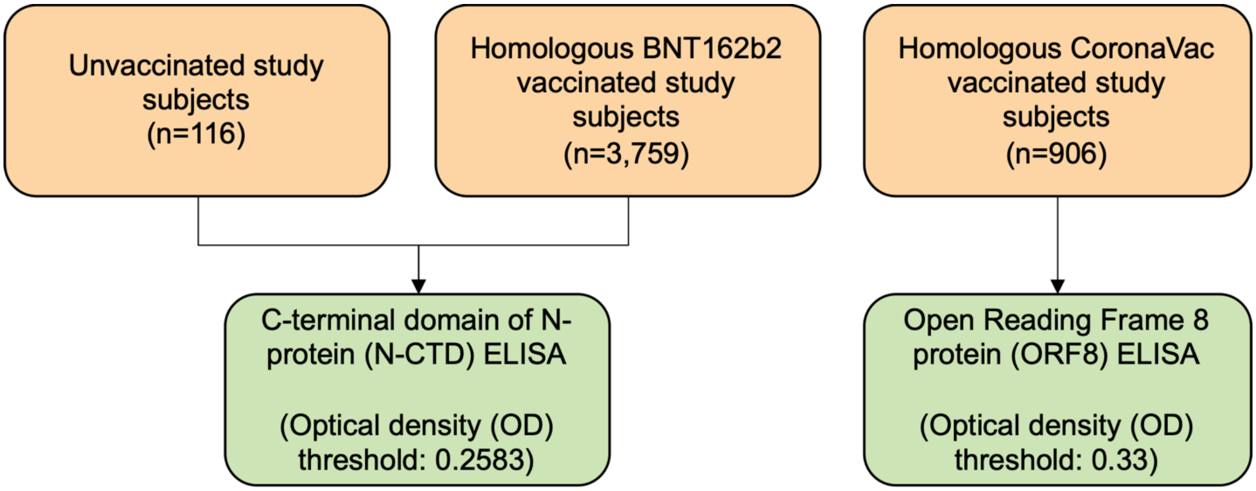


**Extended Data Figure 2: Use of ELISA assay to discriminate infection from vaccine immunity by vaccination cohort.** The above summarises the in-house ELISA assays (green) we used to test for seropositivity amongst study subjects in different vaccination cohorts (orange). We did not test for seropositivity amongst heterologously vaccinated study subjects, study subjects who received vaccines other than BNT162b2 or CoronaVac, or study subjects with an un-determined vaccination history.

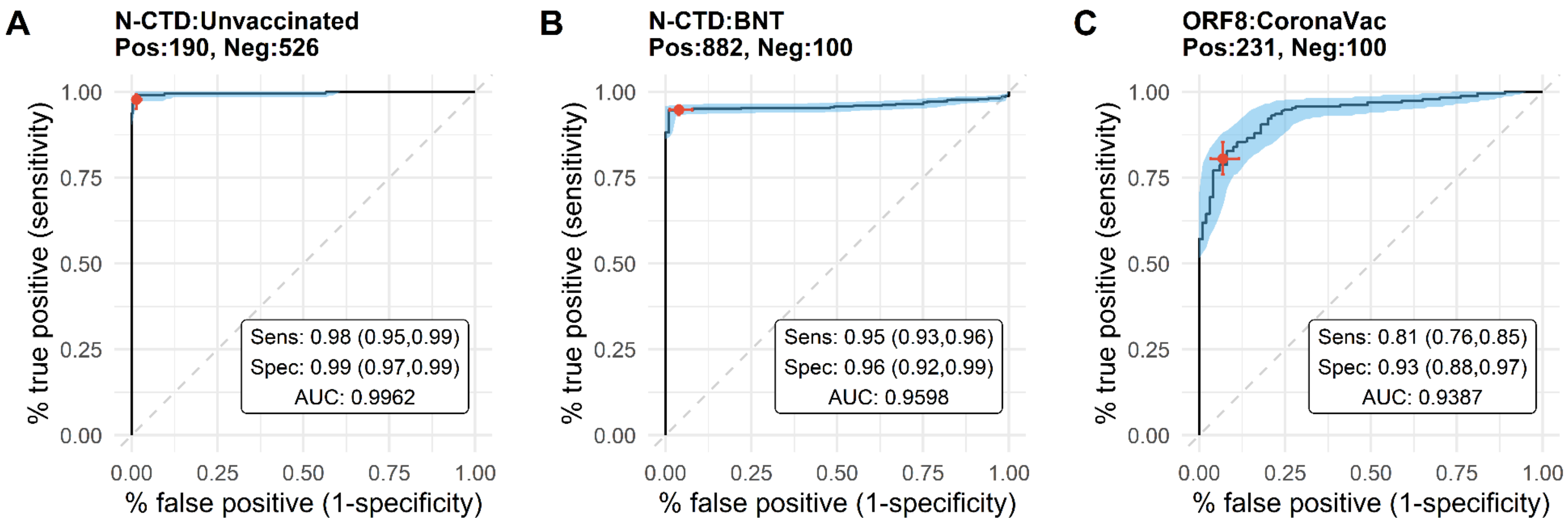


**Extended Data Figure 3: Description of controls, assay performance and Receiver Operating Curves (ROC) by ELISA assay type for detecting Omicron BA.2 infection with in-house ELISA protocols. (A)** N-CTD ELISA amongst unvaccinated controls, tested against 48 unvaccinated study subjects with self-reported infection history, 142 unvaccinated RT-PCR positive convalescent samples ranging from 30 to 401 days after onset of illness and 526 pre-pandemic negative controls; **(B)** N-CTD ELISA amongst controls who were homologously vaccinated with BNT162b2, tested against 881 BNT-vaccinated study subjects with self-reported infection history, 1 BNT-vaccinated RT-PCR positive convalescent sample collected 369 days after onset of illness and 50 (tested twice) non-infected BNT-vaccinated samples collected during 2020-2021, a period of minimal community transmission; and **(C)** ORF8 ELISA amongst controls who were homologously vaccinated with CoronaVac, tested against 231 CoronaVac-vaccinated study subjects with self-reported infection history and 100 non-infected CoronaVac-vaccinated samples collected during 2020-2021, a period of minimal community transmission. Shaded areas indicate 95% confidence region. Red dot and cross represent the median sensitivity and specificity and corresponding confidence intervals of the OD threshold jointly estimated via Gibbs Sampling as described in Statistical Methods above. Pos: positive controls, Neg: negative controls, Sens: sensitivity, Spec: specificity, AUC: area under the curve.

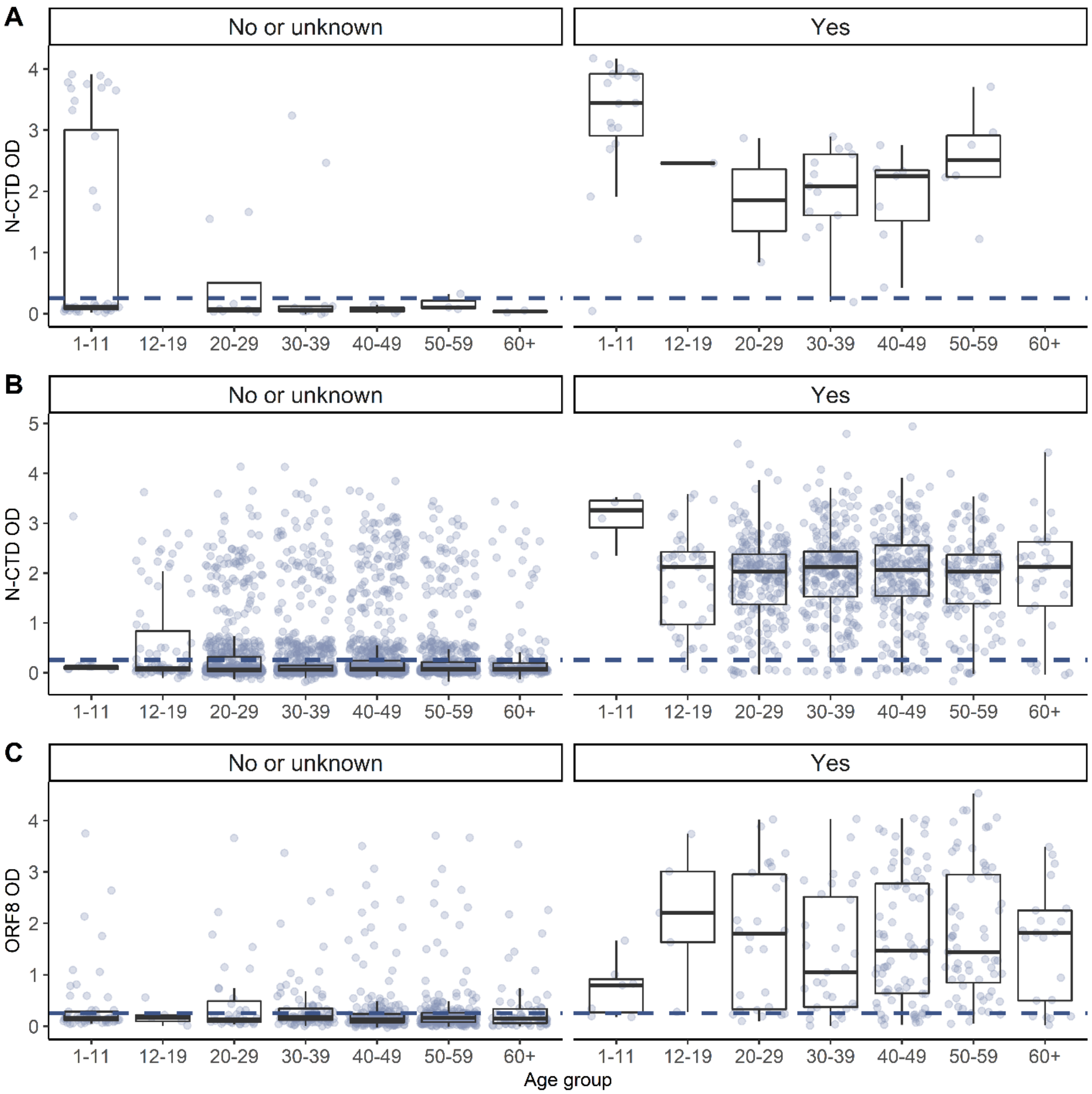


**Extended Data Figure 4: SARS-CoV-2 N-CTD and ORF8 antibody responses amongst study subjects by vaccination history, age and self-reported infection history. A.** Unvaccinated **B.** BNT homologously vaccinated and **C.** CoronaVac homologously vaccinated. Panels on the left correspond to individuals with no self-reported infection history or who did not provide his/her infection history. Panels on the right correspond to individuals with self-reported infection history. The blue dotted line corresponds to the optical density (OD) thresholds for seropositivity (respectively, N-CTD OD of 0.2583 for unvaccinated and BNT homologous vaccinated and ORF8 OD of 0.33 for CoronaVac homologous vaccinated). Each dot represents one study subject. The centre line of each box represents the median, box limits represent the interquartile range (IQR), and the whiskers represent the minimum and maximum observations greater and lesser than the IQR plus 1.5 times IQR respectively.

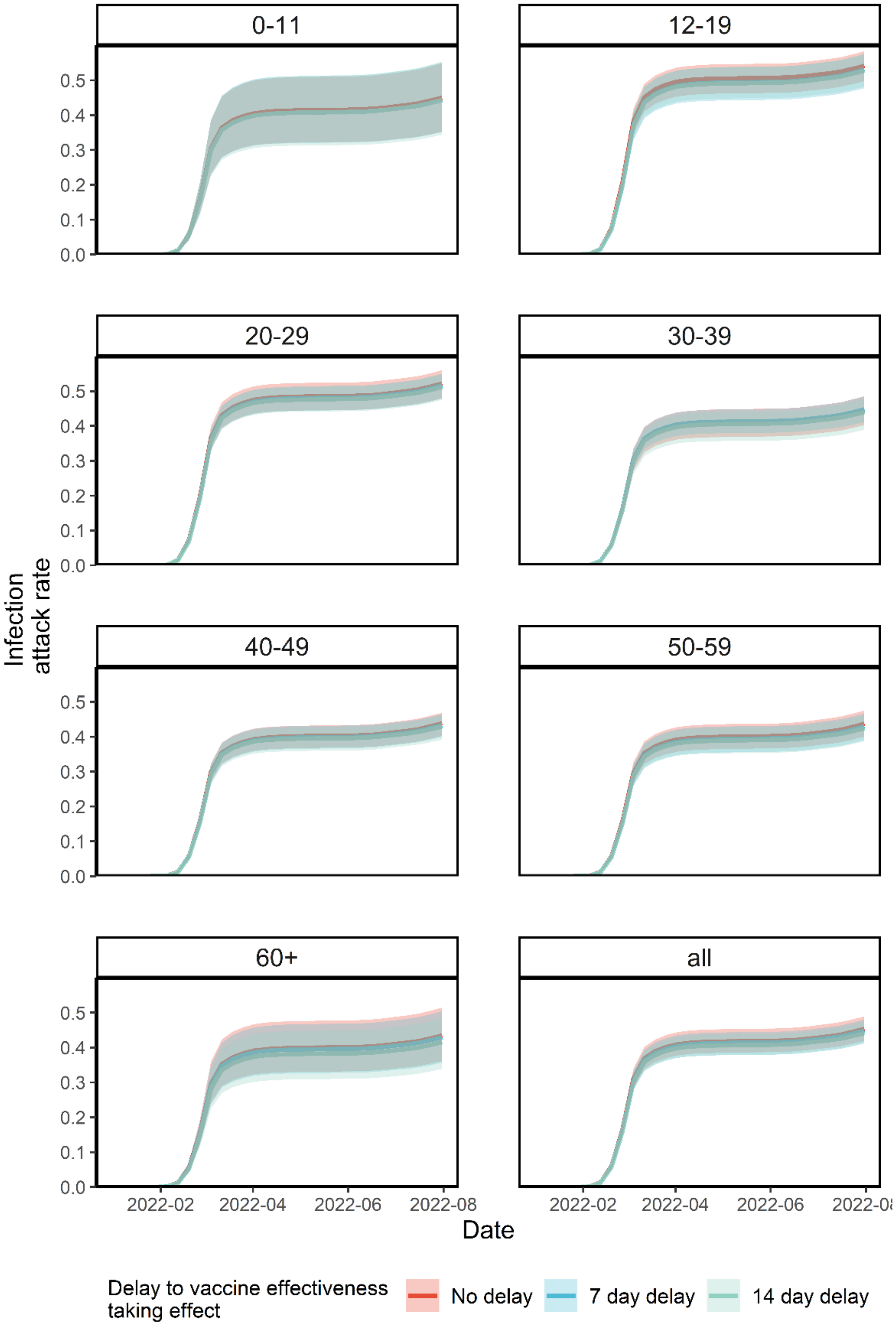


**Extended Data Figure 5: Infection attack rate over time by age group.** IAR estimates among those aged 12-19 or aged 60 or above were less accurate as no subjects were between 12 and 17 years old, and few were above 65 years old. The lines indicate posterior medians and shaded bars indicate 95% credible intervals based on the fitted model.

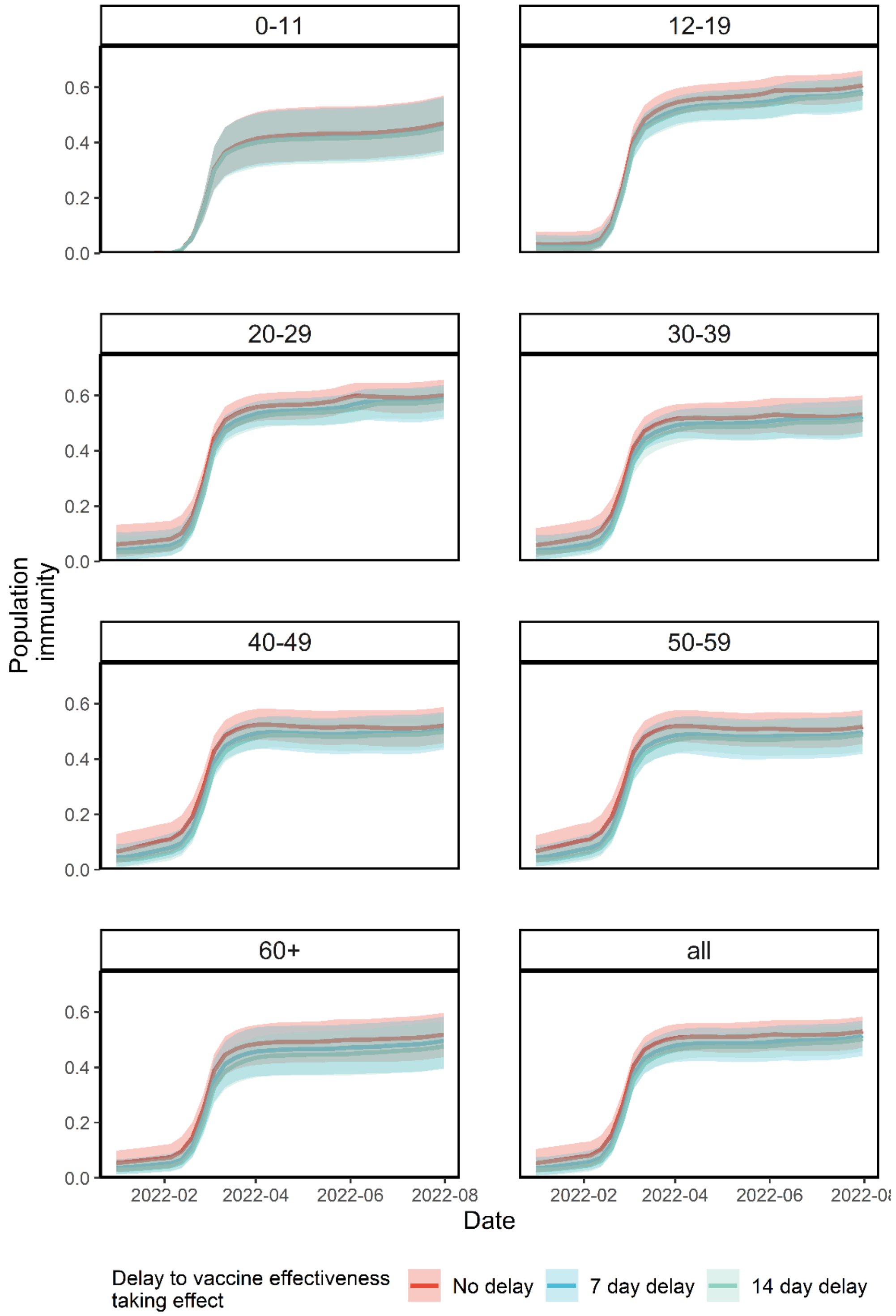


**Extended Data Figure 6: Population immunity over time from vaccination and infection by age group.** Population immunity estimates among those aged 12-19 or aged 60 or above were less accurate as no subjects were between 12 and 17 years old, and few were above 65 years old. The lines indicate posterior medians and shaded bars indicate 95% credible intervals based on the fitted model.

| | Overall (n = 5,242) | | BNT162b2-exclusive (n = 3,759) | | CoronaVac- exclusive (n = 906) | | BNT162b2 and CoronaVac (n = 461) | | Unvaccinated (n=116) | |
|---|---|---|---|---|---|---|---|---|---|---|
| **Age group (years)** | **Number of donors** | **% of total** | **Number of donors** | **% of subtotal** | **Number of donors** | **% of subtotal** | **Number of donors** | **% of subtotal** | **Number of donors** | **% of subtotal** |
| 1-10 | 136 | 2.6 | 13 | 0.3 | 64 | 7.0 | 0 | 0.0 | 59 | 53.2 |
| 18–19 | 134 | 2.6 | 121 | 3.2 | 10 | 1.1 | 2 | 0.4 | 1 | 0.9 |
| 20–29 | 943 | 18.0 | 838 | 22.3 | 61 | 6.7 | 35 | 7.6 | 9 | 8.1 |
| 30–39 | 1183 | 22.6 | 945 | 25.1 | 124 | 13.7 | 91 | 19.7 | 23 | 20.7 |
| 40–49 | 1439 | 27.5 | 1008 | 26.8 | 280 | 30.8 | 143 | 31.0 | 8 | 7.2 |
| 50–59 | 1086 | 20.7 | 651 | 17.3 | 281 | 30.9 | 145 | 31.5 | 9 | 8.1 |
| 60–69 | 321 | 6.1 | 186 | 4.9 | 88 | 9.7 | 45 | 9.8 | 2 | 1.8 |
| **Sex** | | | | | | | | | | |
| Female | 2680 | 51.1 | 1974 | 52.5 | 434 | 47.8 | 216 | 46.9 | 56 | 50.5 |
| Male | 2548 | 48.6 | 1787 | 47.5 | 473 | 52.1 | 245 | 53.1 | 43 | 38.7 |
| Not-determinable | 14 | 0.3 | 1 | 0.0 | 1 | 0.1 | 0 | 0.0 | 12 | 10.8 |
| **Doses received** | | | | | | | | | | |
| 0 | 116 | 2.2 | 0 | 0.0 | 0 | 0.0 | 0 | 0.0 | 116 | 100.0 |
| 1 | 72 | 1.4 | 38 | 1.1 | 34 | 3.8 | 0 | 0.0 | 0 | 0.0 |
| 2 | 1174 | 22.4 | 930 | 24.7 | 223 | 24.6 | 21 | 4.6 | 0 | 0.0 |
| 3 | 3794 | 72.4 | 2760 | 73.4 | 621 | 68.4 | 413 | 89.6 | 0 | 0.0 |
| 4 | 86 | 1.6 | 31 | 0.8 | 28 | 3.1 | 27 | 5.9 | 0 | 0.0 |

**Extended Data Table 1:** Characteristics of study participants, April 2022 – July 2022 (n = 5,310), excluding 67 participants with non-BNT162b2 or non-CoronaVac, or undetermined, vaccination history and 1 participant with undetermined age.
